# The Child Replacement Effect: Shorter Subsequent Birth Intervals Following Early Child Loss in Sub-Saharan Africa

**DOI:** 10.64898/2026.01.15.26344239

**Authors:** Yared Mekonnen

**Affiliations:** Mela Research, PO Box 34422, Addis Ababa, Ethiopia

**Keywords:** Child replacement effect, Birth intervals, Neonatal mortality, Infant mortality, Sub-Saharan Africa

## Abstract

**Background:** Short birth intervals raise maternal, perinatal, and child health risks. Sub-Saharan Africa (SSA) has the world’s highest fertility and elevated child mortality. While short intervals’ effects on child mortality are well established, the reverse link is less studied. The child replacement effect posits that early child death shortens subsequent birth intervals. This study examines and quantifies this association across 26 SSA countries.

**Methods:** This study pooled recent DHS data from 26 SSA countries, included women aged 15–49 with at least one live birth, and analyzed all births and consecutive intervals (closed and right-censored) from birth histories. The primary exposure was prior child’s survival status: survived infancy (reference), neonatal death, or post-neonatal infant death. Outcomes were subsequent birth interval duration (months) and hazard of next birth. This study reported median interval lengths with interquartile range (IQR). Multilevel Weibull parametric survival model with gamma shared frailty were used to estimate adjusted hazard ratios (AHR) with 95% confidence intervals (Cis).

**Result:** The pooled sample from 26 SSAs included 271,117 women contributing 1,181,477 births. Median subsequent birth intervals were 38 months (IQR 26–77) after a child survived to infancy, 24 months (IQR 16–36) after neonatal death, and 25 months (IQR 18–36) after post-neonatal infant death. Replacement intervals clustered tightly around 21–24 months following child loss compared with survival. In fully adjusted multivariate model, prior neonatal death was associated with a 69% higher hazard of next birth (AHR 1.69; 95% CI 1.64–1.74), and post-neonatal infant death with a 50% higher hazard (AHR 1.50; 95% CI 1.46–1.54) versus survival. Country-specific adjusted HR estimates were consistently positive and significant across all 26 countries, ranging from 1.4 to 4.3 for neonatal death and 1.3 to 3.4 for post-neonatal death, with slightly stronger effects after neonatal than post-neonatal infant death.

**Conclusion:** Early child mortality is a strong predictor of shortened subsequent birth intervals throughout SSA, consistent with a persistent child replacement effect. Integrating bereavement-sensitive postpartum family planning into maternal, newborn, and child health programs could promote optimal birth spacing, mitigate replacement-driven accelerations of childbearing, and contribute toward reproductive goals.

## Background

Sub-Saharan Africa (SSA) continues to experience the highest fertility levels worldwide, with a total fertility rate (TFR) of over four children per woman in 2024 [1,2]. This marks a gradual decline from levels exceeding six children per woman in the 1950s [3]. Progress has been driven in part by improvements in female education, urbanization, and family planning access [4]. However, fertility remains elevated and uneven across the region, with high-fertility countries contrasting those with lower rates. Persistent preferences for larger families, limited modern contraceptive use, and cultural factors contribute to sustained high fertility [5]. Birth spacing practices in SSA have traditionally featured prolonged breastfeeding and postpartum abstinence [6]. Recent contraceptive uptake contributes to gradual lengthening of intervals in some areas, alongside declining desired family sizes. Recent Demographic and Health Surveys (DHS) show that short birth to pregnancy intervals (<24 months) affect a substantial proportion of births, particularly in high-fertility contexts, and are associated with adverse health outcomes [7,8]. Trends indicate slight lengthening in Eastern and Southern Africa due to socioeconomic improvements, though short intervals remain prevalent in high-fertility settings [4,8]. These spacing patterns influence overall fertility levels and intersect with child survival outcomes, as short intervals elevate risks of adverse maternal, perinatal and child events [9, 10].

In parallel, Sub-Saharan Africa bears a disproportionate share of global child mortality, accounting for 58% of under-five deaths worldwide in 2023 despite comprising a smaller proportion of the global child population [11]. Although under-five mortality has declined substantially- from over 180 deaths per 1,000 live births in the 1990s to approximately 68 per 1,000 in 2023- rates remain elevated in numerous countries [11,12]. A substantial proportion of these deaths occur in the early periods of life, with neonatal mortality estimated at around 26–27 per 1,000 live births, and early neonatal mortality accounting for approximately three-quarters of neonatal deaths [12,13]. The infant mortality rate in Sub-Saharan Africa stands at around 44 per 1,000 live births in 2023, highlighting the persistent vulnerability of newborns and young infants in the region [11]. Progress in reducing neonatal and infant mortality has been slower than for older children, with neonatal deaths comprising about 40% of under-five deaths in the region, underscoring the need for targeted interventions in the perinatal and postnatal periods [14].

The child replacement effect, as formalized by Preston (1978), posits that the death of a child prompts parents to accelerate subsequent childbearing through shortened birth intervals or additional births in an effort to replace the loss and achieve their desired number of surviving offspring [15]. The effects of infant and child mortality on birth intervals operate through both physiological mechanisms and volitional behaviors. The most prominent physiological mechanism is lactational amenorrhea, whereby breastfeeding suppresses ovulation after a birth; when a child dies, breastfeeding typically ends, allowing ovulation to return more quickly. The volitional replacement effect, also called the behavioral replacement effect, refers to the intentional response by parents who conceive earlier or have additional children after a child’s death to replace the lost child and attain their target family size or number of surviving offspring [16]. A related insurance effect, sometimes termed hoarding, involves precautionary extra births in anticipation of possible future child losses [15–17].

Research on the child replacement effect in sub-Saharan Africa remains limited. Nevertheless, a small number of studies have documented its presence in specific settings, including rural Tanzania [18], Cameroon [17], and Ghana and Kenya [19]. These studies generally indicate that child mortality is associated with shorter subsequent birth intervals and accelerated progression to higher parities, through a combination of physiological and volitional mechanisms.

Amid ongoing improvements in child survival, socioeconomic development, and access to family planning across the region, understanding these replacement dynamics offers valuable insight into a key factor that continues to sustain persistently high fertility levels. The present study addresses important gaps in the literature by pooling recent DHS data from 26 SSA countries to examine the association between prior neonatal or post-neonatal infant death and subsequent birth intervals.

## Methods

### Data source and study sample

This analysis used Demographic and Health Surveys (DHS), large nationally representative household surveys in low- and middle-income countries that collect population, health, and nutrition data, primarily funded by USAID [19]. We included the most recent DHS from 26 sub-Saharan African countries (2013–2024), selected based on availability of individual-level records and complete birth histories. Standardized DHS questionnaires and methods enabled pooling across countries.

The study population comprised women aged 15–49 years with at least one live birth. We analyzed all births and consecutive birth intervals from each woman’s full birth history, including closed intervals and the open interval (from the most recent birth to interview, right-censored).

### Study variables

The outcome was the duration of each eligible birth interval (in months) from the birth of the preceding child to either the next birth (closed interval) or the interview date (open interval). The primary exposure, previous child mortality, categorized as survived to infancy ( ≥12 months), neonatal death, or post-neonatal infant death based on the child at the interval start. Neonatal death was defined as death within the first month of life (<1 month) and post-neonatal infant death as deaths between 1 and 11 months [20].

Covariates included maternal age at the previous birth (<20 years, 20–24, 25–29, 30–34, 35-49 years), multiple birth status at interval start (singleton vs. multiple), sex of the preceding child (female vs. male), residence (rural vs. urban), current marital status (never married, currently married/living with partner, ever married but divorced/widowed/separated), maternal education (no education, primary, secondary or higher), household wealth quintile, as provided in the DHS dataset (poorest to richest), number of living children immediately before the interval start (0–2, 3–4, ≥5), prior sex composition of living children (balanced or one child, more boys, more girls), year of birth of the child at interval start (continuous), and country fixed effects (dummy variables for unobserved country-level heterogeneity).

### Statistical analysis

This study employed a multilevel Weibull parametric survival model with gamma shared frailty to examine the association between child loss and subsequent birth interval, while accounting for clustering of children within mothers and the complex survey design of the data. The model included a vector of fixed-effect covariates comprising the categorical indicator of previous child death experience together with other relevant covariates. Child-level probability weights were applied to adjust for unequal selection probabilities, non-response, and post-stratification.

The Weibull hazard function for child *i* of mother *j* at time *t* is expressed as:

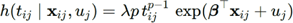

Where;

- t_ij_ is the age in months at death or censoring for child *i* of mother *j*,
- h_0_(t)=λpt^p−1^ is the Weibull baseline hazard function with scale parameter λ>0 and shape parameter p>0,
- x_ij_ is the vector of observed covariates,
- β is the corresponding vector of fixed-effect regression coefficients,
- u_j_ ∼ N(0, σ_u_^2^) is the random intercept (frailty term) at the maternal level, assumed to be normally distributed on the log-hazard scale.

Two multivariate models are presented: Model 1 was partially adjusted and included the primary exposure variable, previous child mortality (categorized as neonatal death, post-neonatal infant death, or survival to infancy), along with controls for country (fixed effects) and year of birth. Model 2 was a fully adjusted model that additionally incorporated the following covariates: maternal age at the previous birth, multiple birth status of the previous child, sex of the previous child, residence, marital status, maternal education, household wealth quintile, number of living children, and prior sex composition of children. Adjusted hazard ratios (AHRs) were reported with 95% confidence intervals and p-values.

Data management and statistical analyses were conducted using Stata version 18 (StataCorp, College Station, TX, USA). Missing data were minimal, owing to the rigorous data collection protocols employed by the DHS. Observations with missing values on key indicators were excluded from the analysis, as these cases represented less than 2% of the sample. All estimates were weighted using the DHS survey weights to adjust for the complex sampling design and unequal selection probabilities.

## Results

### Sample distribution of women, births, and neonatal/infant deaths by country

Table 1 describes the analytic sample, which comprises 271,117 women aged 15–49 who had at least one live birth and contributed a total of 1,181,477 births. These data are drawn from the most recent DHS conducted between 2013 and 2024 in 26 SSA countries. Among these women, the pooled mean number of births per woman was 4.4, with substantial country-level variation ranging from 2.9 in Zimbabwe to 5.4 in Chad.

**Table 1.** Descriptive characteristics of women aged 15–49 with at least one live birth: Number of births, average number of births, and early child mortality (neonatal and post-neonatal infant deaths), pooled and by country, Demographic and Health Surveys, SSA, 2013–2024

| Country | Survey year | Number of women (with at least one live birth) | Number of births | Average number of births per woman | Neonatal deaths (%) | Post-neonatal infant deaths (%) |
| --- | --- | --- | --- | --- | --- | --- |
| Pooled (total) | 2013-2024 | 271,117 | 1,181,477 | 4.4 | 2.71 | 2.65 |
| Angola | 2023-2024 | 8,995 | 37,767 | 4.2 | 2.89 | 3.38 |
| Benin | 2017-2018 | 9,333 | 45,853 | 4.9 | 2.93 | 2.41 |
| Burundi | 2016-2017 | 11,014 | 45,419 | 4.1 | 2.65 | 3.38 |
| Burkina Faso | 2021 | 10,300 | 48,745 | 4.7 | 2.27 | 1.95 |
| Cameroon | 2018-2019 | 9,048 | 33,988 | 3.8 | 2.61 | 2.34 |
| Chad | 2014-2015 | 12,838 | 68,989 | 5.4 | 3.15 | 4.01 |
| Congo-DRC | 2023-2024 | 17,509 | 80,890 | 4.6 | 2.76 | 3.65 |
| Côte d'Ivoire | 2021 | 10,942 | 39,979 | 3.7 | 2.54 | 2.25 |
| Ethiopia | 2016 | 9,357 | 41,392 | 4.4 | 3.85 | 2.80 |
| Gabon | 2020-2021 | 4,621 | 23,232 | 5.0 | 1.95 | 1.26 |
| Gambia | 2019-2020 | 6,270 | 31,665 | 5.1 | 2.85 | 1.68 |
| Ghana | 2022-2023 | 8,326 | 34,663 | 4.2 | 2.30 | 1.36 |
| Guinea | 2018 | 5,801 | 28,887 | 5.0 | 2.88 | 2.54 |
| Kenya | 2022 | 20,537 | 77,381 | 3.8 | 1.91 | 1.43 |
| Liberia | 2019-2020 | 5,549 | 24,765 | 4.5 | 3.56 | 4.01 |
| Malawi | 2015-2016 | 16,984 | 68,074 | 4.0 | 2.54 | 2.66 |
| Mali | 2023-2024 | 10,267 | 53,413 | 5.2 | 3.38 | 2.70 |
| Nigeria | 2023-2024 | 21,624 | 104,557 | 4.8 | 2.90 | 2.00 |
| Rwanda | 2019-2020 | 7,791 | 30,820 | 4.0 | 3.13 | 3.65 |
| Senegal | 2023 | 8,464 | 39,490 | 4.7 | 2.30 | 1.23 |
| Sierra Leone | 2019 | 11,236 | 40,543 | 3.6 | 2.79 | 4.65 |
| Tanzania | 2022 | 8,412 | 40,394 | 4.8 | 2.54 | 3.00 |
| Togo | 2013-2014 | 5,785 | 26,264 | 4.5 | 2.77 | 2.85 |
| Uganda | 2016 | 13,745 | 57,906 | 4.2 | 2.42 | 3.07 |
| Zambia | 2024 | 9,116 | 35,610 | 3.9 | 2.36 | 2.39 |
| Zimbabwe | 2015 | 7,253 | 20,791 | 2.9 | 2.26 | 2.02 |

Prior child mortality served as the primary exposure and was measured among birth intervals following a previous live birth. In the pooled sample, 2.71% of prior children died in the neonatal period and 2.65% died in the post-neonatal infant period (1–11 months). Neonatal death rates peaked in Ethiopia (3.85%) and Liberia (3.56%), while post-neonatal rates were highest in Sierra Leone (4.65%) and Chad (4.01%). Lower rates appeared in Kenya (1.91% neonatal and 1.43% post-neonatal), Ghana (2.30% and 1.36%), Gabon (1.95% and 1.26%), and Senegal (2.30% and 1.23%).

This substantial heterogeneity in the distribution of live births and early child survival, likely driven by differences in survey timing, socioeconomic conditions, and health systems, provides essential variation for testing the child replacement hypothesis.

### Descriptive analysis of birth intervals by child survival status

Table 2 presents median birth intervals in months (with interquartile ranges) stratified by the survival status of the previous child, for the pooled sample and individually for each of the 26 SSA countries. Across the pooled sample of 1,181,477 births, the overall median birth interval was 37 months (IQR: 25–72). When the previous child survived, the median interval was 38 months (IQR: 26–77). Following a neonatal death, the median dropped sharply to 24 months (IQR: 16–36), and following a post-neonatal infant death to 25 months (IQR: 18–36). This represents a reduction of 14 months for neonatal deaths and 13 months for post-neonatal deaths compared to intervals after a surviving child.

**Table 2.** Median birth interval in months and interquartile range (IQR) by previous child mortality status, pooled and by country, Demographic and Health Surveys, SSA, 2013–2024

| Country | Survived<br>previous child<br>Median (IQR) | Previous<br>Neonatal death<br>Median (IQR) | Previous<br>Infant death<br>Median (IQR) | Overall<br>Median (IQR) | Number of<br>children<br>(complete birth<br>history) |
| --- | --- | --- | --- | --- | --- |
| **Pooled (total)** | 38 (26–77) | 24 (16–36) | 25 (18–36) | 37 (25–72) | 1,142,913 |
| Angola | 36 (25–71) | 23 (15–34) | 24 (17–36) | 35 (25–66) | 42,002 |
| Benin | 37 (27–65) | 26 (17–38) | 26 (19–35) | 36 (26–62) | 45,853 |
| Burundi | 35 (26–59) | 21 (14–29) | 23 (17–33) | 34 (25–55) | 45,419 |
| Burkina Faso | 41 (29–76) | 25 (17–39) | 27 (19–43) | 41 (28–73) | 48,745 |
| Cameroon | 35 (25–74) | 23 (16–31) | 25 (18–32) | 34 (24–69) | 33,988 |
| Chad | 29 (22–46) | 23 (16–31) | 25 (18–32) | 28 (22–45) | 68,989 |
| Congo - DRC | 34 (24–61) | 24 (16–35) | 24 (17–35) | 33 (24–57) | 59,276 |
| Côte d'Ivoire | 43 (28–92) | 26 (17–38) | 26 (19–40) | 42 (27–86) | 39,979 |
| Ethiopia | 35 (24–63) | 23 (14–33) | 22 (15–33) | 34 (24–60) | 41,392 |
| Gabon | 49 (28–133) | 35 (21–64) | 38 (21–67) | 48 (28–131) | 23,232 |
| Gambia | 37 (27–68) | 25 (17–35) | 26 (18–38) | 36 (27–66) | 31,665 |
| Ghana | 47 (30–110) | 28 (18–45) | 28 (19–45) | 46 (29–103) | 34,663 |
| Guinea | 38 (27–72) | 26 (20–38) | 27 (21–37) | 37 (26–69) | 28,887 |
| Kenya | 50 (28–139) | 24 (15–41) | 26 (17–41) | 48 (28–226) | 77,381 |
| Liberia | 44 (28–109) | 26 (17–41) | 25 (18–36) | 42 (27–97) | 24,765 |
| Malawi | 41 (28–81) | 23 (15–35) | 25 (17–35) | 40 (27–76) | 68,074 |
| Mali | 33 (24–59) | 24 (18–34) | 25 (19–34) | 32 (24–57) | 33,379 |
| Nigeria | 34 (24–64) | 23 (16–33) | 24 (18–34) | 34 (24–61) | 104,557 |
| Rwanda | 38 (26–79) | 24 (15–44) | 27 (19–43) | 37 (26–74) | 30,058 |
| Senegal | 39 (26–76) | 27 (19–45) | 30 (21–50) | 38 (26–74) | 39,490 |
| Sierra Leone | 41 (27–92) | 25 (18–37) | 25 (19–36) | 39 (26–82) | 40,543 |
| Togo | 38 (27–81) | 24 (16–37) | 25 (18–37) | 37 (26–76) | 37,169 |
| Uganda | 40 (28–76) | 28 (19–40) | 29 (21–43) | 39 (28–73) | 26,264 |
| Niger | 33 (24–64) | 20 (13–29) | 23 (16–32) | 32 (23–60) | 57,906 |
| Tanzania | 39 (28–80) | 27 (18–40) | 28 (19–42) | 39 (27–76) | 38,446 |
| Zimbabwe | 54 (35–139) | 22 (14–36) | 24 (16–34) | 52 (33–156) | 20,791 |

This pattern of substantially shorter birth intervals following child mortality was observed consistently across all 26 countries, with no exceptions. Among intervals following the survival of the previous child, median lengths varied widely, from a low of 29 months in Chad to a high of 54 months in Zimbabwe, reflecting diverse fertility behaviors, socioeconomic conditions, and levels of contraceptive access across countries. In contrast, median intervals following a neonatal or post-neonatal infant death showed striking consistency, generally ranging between 22 and 28 months across countries, with only a few outliers.

Country-specific results further illustrate the robustness of the association. In countries with longer typical spacing when the previous child survived, the absolute reduction after child loss was particularly pronounced. For example, Zimbabwe had the longest median interval after survival (54 months, IQR: 35–139) but one of the shortest after neonatal death (22 months, IQR: 14–36), yielding a 32-month difference. Kenya showed a similar large gap (50 months after survival vs. 24 months after neonatal death; difference of 26 months) and Gabon (49 vs. 35 months; 14 months). Ghana (47 vs. 28 months) and Côte d’Ivoire (43 vs. 26 months) also exhibited differences exceeding 15 months. In countries with shorter baseline spacing, the absolute differences were smaller but still substantial. Chad, with the shortest median after survival (29 months, IQR: 22–46), had medians of 23 months after neonatal and 25 months after infant death (differences of 6 and 4 months). Niger displayed a comparable pattern (33 vs. 20 months after neonatal death; 13-month difference). Mali (33 vs. 24–25 months) and the Democratic Republic of the Congo (34 vs. 24 months) similarly showed reductions of 9–10 months.

In many countries, median intervals following neonatal and post-neonatal infant deaths were nearly identical, suggesting similar reproductive responses regardless of whether the loss occurred in the first month or later in infancy.

The interquartile ranges (IQRs) highlight substantial heterogeneity in birth spacing behavior. Following a child’s survival, intervals exhibited much wider variation (pooled IQR: 26–77 months), reflecting diverse spacing patterns in the absence of a replacement motive. In contrast, intervals following child mortality were consistently narrower (pooled IQR: 16–36 months). This pattern holds across all countries in the study, indicating that replacement births tend to cluster within a relatively short and predictable postpartum window, even in highly diverse settings.

### Frequency distributions of birth intervals by child survival status

Figure 1 presents density plots of subsequent birth interval lengths (in months) stratified by the survival status of the previous child in the pooled sample: survived infancy, neonatal death, and post-neonatal infant death. The distribution following survival of the previous child to infancy is broad and right-skewed and a gradual decline extending far into the tail beyond 150 months. This wide spread reflects substantial heterogeneity in birth spacing when the previous child survives, influenced by varying durations of breastfeeding, postpartum amenorrhea, contraceptive use, and intentional family planning across households and countries. In stark contrast, the distributions following neonatal and post-neonatal infant death are markedly shifted leftward, with much higher peak densities and considerably narrower spreads. Both curves rise sharply to a prominent peak around 22–25 months, reaching densities higher than the peak for surviving children. The neonatal and post-neonatal lines are nearly superimposed for the initial rise and peak, indicating almost identical timing of subsequent births regardless of whether the previous child died in the neonatal period or later in infancy. After the peak, both decline rapidly, with very little mass beyond 80-90 months, demonstrating that replacement births following early child loss are highly concentrated in a short postpartum window. These density distributions provide visual evidence supporting the median differences reported in Table 2 and highlight both the magnitude of interval shortening after child loss and the reduced variability in spacing behavior under replacement conditions.

**Figure 1.**
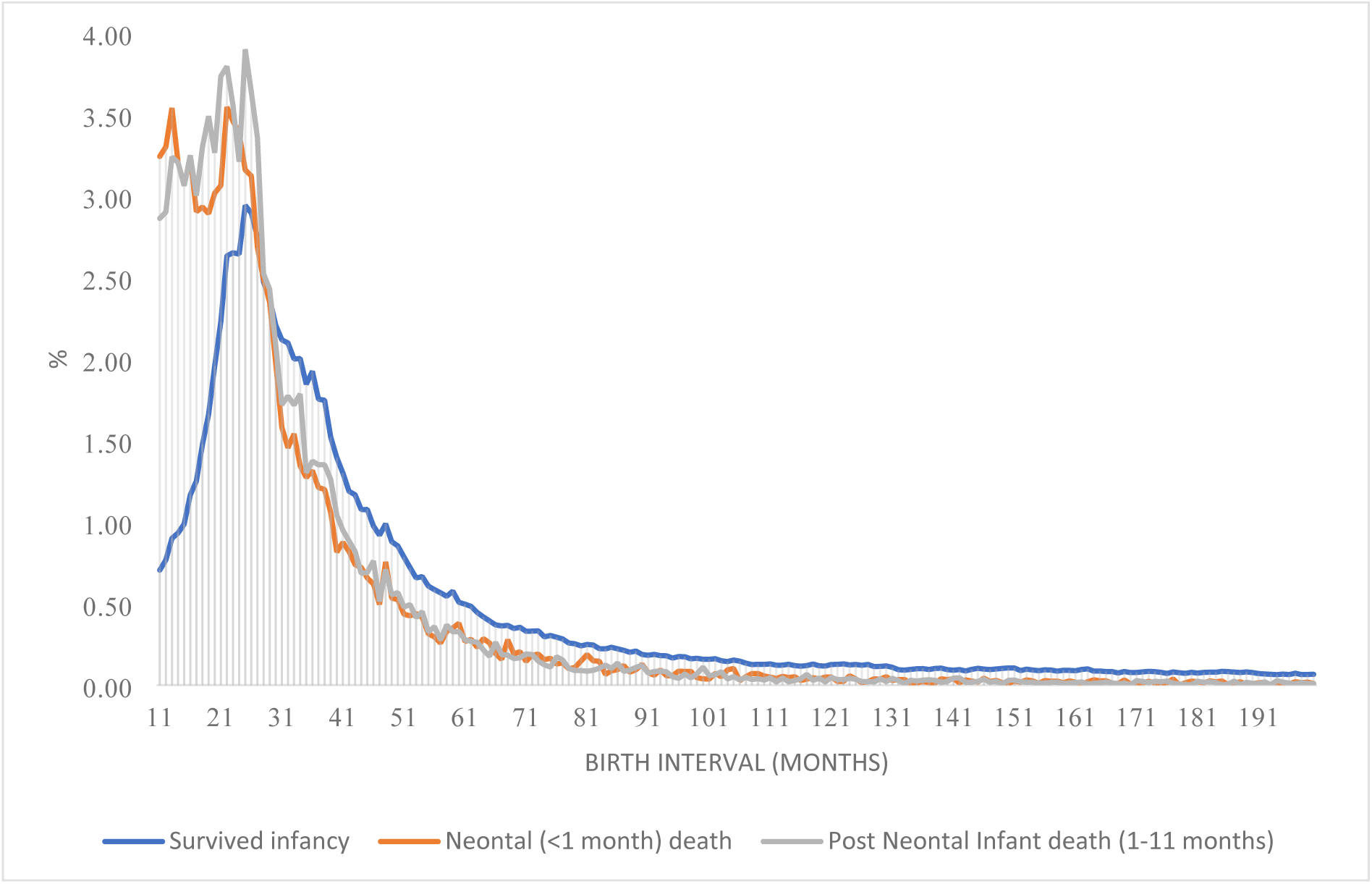
Frequency distributions (pooled) of subsequent birth intervals by previous child survival status, Demographic and Health Surveys, SSA, 2013–2024

### Multivariate results on the pooled sample

In multivariable analyses (Table 3), previous child mortality was strongly associated with shorter subsequent birth intervals, even after full adjustment. In the minimally adjusted Model 1, neonatal death and post-neonatal infant death were associated with 2.40 (95% CI: 2.35-2.45) and 2.47 (95% CI: 2.42-2.51) times higher hazard of shorter intervals, respectively (both P < 0.001). These associations remained highly significant but with reduced effect sizes in the fully adjusted Model 2 (AHR 1.69, 95% CI: 1.64-1.74 for neonatal death; AHR 1.50, 95% CI: 1.46-1.54 for post-neonatal infant death; both P < 0.001). Thus, even after extensive confounding control and accounting for unobserved maternal heterogeneity via frailty, women who experienced a previous neonatal death had a 69% higher hazard of a subsequent birth, while those experiencing a post-neonatal infant death had a 50% higher hazard, compared to women whose previous child survived. The slightly stronger adjusted effect for neonatal versus post-neonatal death may reflect shorter breastfeeding duration and earlier return of fecundity when death occurs very early.

**Table 3.** Adjusted hazard ratios (AHR) and 95% Confidence Intervals (Cis) from multilevel Weibull parametric survival model with gamma shared frailty models for the effect of previous child mortality on subsequent birth intervals, adjusted for selected covariates, Demographic and Health Surveys, SSA, 2013–2024

| Variables | Model 1 |  |  | Model 2 |  |  |
| --- | --- | --- | --- | --- | --- | --- |
|  | AHR | 95% CI | P-value | AHR | 95% CI | P-value |
| **Previous child mortality (Ref: survived)** |  |  |  |  |  |  |
| Neonatal death | 2.40 | 2.35-2.45 | <0.001 | 1.69 | 1.64-1.74 | <0.001 |
| Post-neonatal infant death | 2.47 | 2.42-2.51 | <0.001 | 1.50 | 1.46-1.54 | <0.001 |
| **Maternal age at previous birth (Ref: <20 years)** |  |  |  |  |  |  |
| 20–24 years |  |  |  | 1.07 | 1.05-1.08 | <0.001 |
| 25–29 years |  |  |  | 1.11 | 1.09-1.13 | <0.001 |
| 30–34 years |  |  |  | 1.15 | 1.12-1.18 | <0.001 |
| ≥35 years |  |  |  | 1.05 | 1.01-1.10 | 0.008 |
| **Previous interval was multiple birth (Ref: No)** |  |  |  |  |  |  |
| Yes |  |  |  | 0.85 | 0.83-0.88 | <0.001 |
| **Previous child male (Ref: female)** |  |  |  |  |  |  |
| Male |  |  |  | 1.00 | 0.99-1.01 | 0.492 |
| **Rural residence (Ref: urban)** |  |  |  |  |  |  |
| Rural |  |  |  | 1.08 | 1.06-1.09 | <0.001 |
| **Marital status (Ref: never married)** |  |  |  |  |  |  |
| Currently married/living with partner |  |  |  | 1.27 | 1.21-1.34 | <0.001 |
| Ever married (divorced/widowed/separated) |  |  |  | 0.98 | 0.93-1.04 | 0.576 |
| **Maternal education (Ref: no education)** |  |  |  |  |  |  |
| Primary |  |  |  | 1.01 | 0.97-1.02 | 0.139 |
| Secondary or higher |  |  |  | 0.96 | 0.95-0.98 | <0.001 |
| **Household wealth quintile (Ref: poorest)** |  |  |  |  |  |  |
| Poorer |  |  |  | 0.93 | 0.92–0.95 | <0.001 |
| Middle |  |  |  | 0.87 | 0.86-0.89 | <0.001 |
| Richer |  |  |  | 0.82 | 0.80-0.83 | <0.001 |
| Richest |  |  |  | 0.70 | 0.68-0.72 | <0.001 |
| **Number of living children before index birth (Ref: 0–2)** |  |  |  |  |  |  |
| 3–4 |  |  |  | 1.05 | 1.04-1.06 | <0.001 |
| ≥5 |  |  |  | 1.17 | 1.15-1.19 | <0.001 |
| **Prior sex composition (Ref: balanced or only one child)** |  |  |  |  |  |  |
| More boys than girls |  |  |  | 1.01 | 0.99-1.02 | 0.347 |
| More girls than boys |  |  |  | 1.02 | 1.00-1.03 | 0.021 |
| **Year of birth (an increase of one year)** | 0.96 | 0.95-0.97 | <0.001 | 0.96 | 0.96-0.97 | <0.001 |
| Both Models 1 and 2 are adjusted for country-level fixed effects and maternal-level shared gamma frailty (random effects)<br>AHR=Adjusted Hazard Ratio; CI=Confidence Interval; Ref=Reference category |  |  |  |  |  |  |

Several covariates in Model 2 were independently associated with birth interval length. Older maternal age at the previous birth was generally linked to higher hazards (shorter intervals) compared to age <20 years, peaking at ages 30–34 (AHR 1.15, 95% CI: 1.12-1.18) before declining slightly at ≥35 years (AHR 1.05, 95% CI: 1.01–1.10). Multiple births at the interval start were associated with longer subsequent intervals (AHR 0.85, 95% CI: 0.83-0.88). Rural residence (AHR 1.08, 95% CI: 1.06-1.09) and current marriage/cohabitation (AHR 1.27, 95% CI: 1.21-1.34) predicted shorter intervals. Higher maternal education (secondary or above: AHR 0.96, 95% CI: 0.95–0.98) and greater household wealth (progressively lower AHRs, reaching 0.70 for richest quintile, 95% CI: 0.68-0.72) were associated with longer intervals, consistent with socioeconomic gradients in contraceptive access and desired family size. Higher parity (≥5 living children: AHR 1.17, 95% CI: 1.15-1.19) and a slight preference effect for more girls than boys (AHR 1.02, 95% CI: 1.00-1.03) predicted shorter intervals. A secular trend was observed, with each one-year increase in the year of birth of a child associated with a 4% lower hazard of shorter subsequent birth intervals (AHR 0.96, 95% CI: 0.95–0.97 in both Model 1 and Model 2; both P < 0.001). This indicates a modest temporal shift toward longer birth spacing, independent of other covariates.

### Country-specific multivariate results

Figures 2 and 3 display country-specific multivariate AHRs with 95% CIs in the estimation of the association between child mortality and subsequent birth intervals. These estimates are fully adjusted for all covariates (maternal age at previous birth, multiple birth status, sex of previous child, rural/urban residence, marital status, maternal education, household wealth quintile, number of living children before the interval start, prior sex composition, year of birth of the child at interval start) as well as maternal-level shared frailty to account for unobserved maternal heterogeneity. Figure 2 displays the association following a neonatal death (relative to survival through infancy), while Figure 3 presents the association following a post-neonatal infant death.

**Figure 2.**
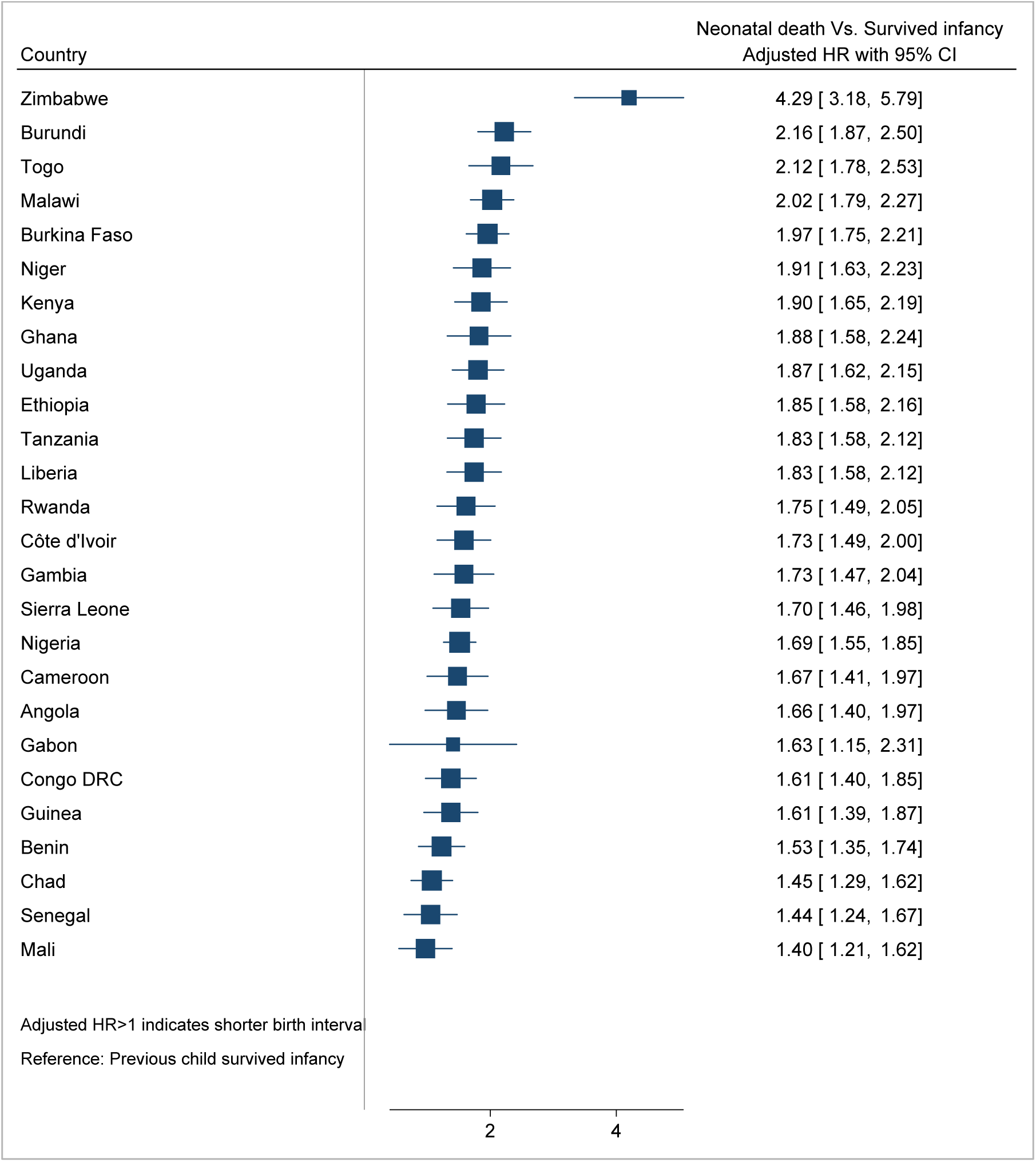
Adjusted hazard ratios (AHR) and 95% Confidence Intervals (Cis) from multilevel Weibull parametric survival model with gamma shared frailty models for the effect of previous neonatal death on subsequent birth intervals, adjusted for selected covariates, by country, Demographic and Health Surveys, SSA, 2013–2024

**Figure 3.**
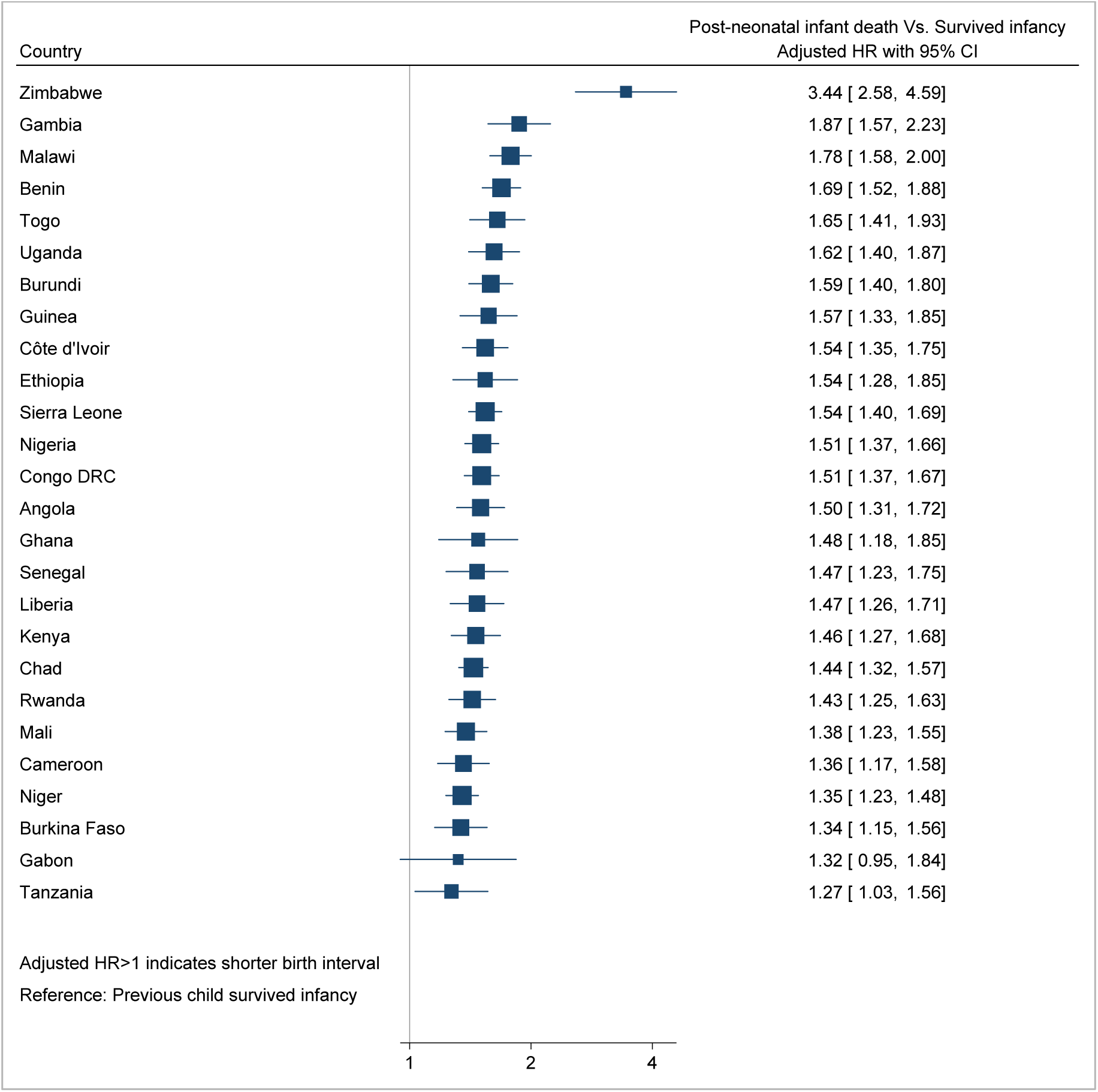
Adjusted hazard ratios (AHR) and 95% Confidence Intervals (Cis) from multilevel Weibull parametric survival model with gamma shared frailty models for the effect of previous post-neonate infant death on subsequent birth intervals, adjusted for selected covariates, by country, Demographic and Health Surveys, SSA, 2013–2024

Across all 26 countries, previous child mortality was consistently and significantly associated with a higher hazard of a subsequent birth (shorter birth intervals), with AHRs greater than 1 and 95% confidence intervals excluding 1 in every country. For neonatal death (Figure 2), fully adjusted AHRs ranged from approximately 1.3 to over 2.2. The strongest associations were observed in Zimbabwe, Kenya, and Gabon (AHRs >2.0), indicating more than a doubling of the hazard of a next birth after neonatal loss even after adjustment for all covariates. Moderately strong effects (AHRs 1.7-2.0) were seen in countries such as Ghana, Côte d’Ivoire, and Liberia. The smallest (yet still highly significant) associations occurred in Chad, Niger, and Mali (AHRs 1.3-1.5).

The pattern for post-neonatal infant death (Figure 3) was similar in direction and significance but with somewhat lower magnitudes in most countries, yielding fully adjusted AHRs ranging from about 1.2 to 1.9. The highest estimates again appeared in Zimbabwe, Kenya, and Gabon (approaching or exceeding 1.8), while the lowest were in Chad, Mali, and Niger (1.2-1.4). In several countries, the adjusted AHRs for neonatal and post-neonatal death were comparable, reflecting the similarity in median intervals observed descriptively.

## Discussion

This multi-country analysis demonstrates a persistent replacement effect across 26 SSA countries spanning diverse fertility and mortality contexts. Early child mortality, particularly neonatal and post-neonatal infant mortality, was associated with markedly shorter subsequent birth intervals in every country examined, with pooled median reductions of 13-14 months and adjusted hazards of shorter birth intervals 50-69% higher. These findings are consistent with intertwined physiological and behavioral pathways, as outlined by Preston [15], whereby child death interrupts lactation (shortening postpartum amenorrhea) and prompts volitional replacement strategies to compensate for lost offspring. The pooled replacement birth intervals clustered around 24-25 months postpartum, suggesting a prominent physiological component in settings with prolonged breastfeeding norms. This timing is well below the WHO-recommended birth-to-birth interval of about 33 months, which is advised to reduce risks of preterm birth, low birth weight, small-for-gestational-age, and maternal complications such as anemia and adverse outcomes [21].

The country-specific, fully adjusted results also confirm that the association between previous child mortality and substantially shorter subsequent birth intervals persists across the diverse contexts of sub-Saharan Africa, even after comprehensive control for covariates as well as intra-woman correlation through shared frailty. The universal positive direction and statistical significance across countries highlight the robust role of child replacement behavior and physiological mechanisms in driving short birth interval following early child loss in the region.

Our findings align with previous studies, though limited in number, that provide evidence of child mortality accelerating subsequent fertility in sub-Saharan Africa, primarily through replacement effects that shorten birth intervals and increase parity progression. In rural Tanzania, Baynes et al. demonstrated strong replacement responses, with hazard ratios for next conception ranging from 3.89 to 4.69 following a child’s death in the interval, resulting in mean interbirth intervals shortening by approximately 8.5 months (from 37.3 to 28.8 months) [18]. This aligns with earlier micro-level findings from Cameroon, where Kuate Defo reported median interval reductions of 5–7 months (up to 6–9 months for neonatal deaths) and short-term conception hazard increases of 61–131%, driven by both physiological mechanisms and volitional replacement [17]. Gyimah and Fernando’s analysis in Ghana and Kenya further supports volitional intent, showing 32–60% elevated odds of higher-parity births after child loss, implying compressed intervals through deliberate acceleration [19]. Collectively, these studies reinforce the enduring influence of child mortality on fertility dynamics in the region.

Our analysis revealed a greater reduction in birth intervals following child loss than previously reported [17.18], demonstrating larger and more consistent effects across diverse SSA countries with varying fertility and mortality levels. The observed strength of the replacement effect in the present study versus prior studies likely reflect several factors. Our analysis leverages complete birth histories (including all closed and open intervals), a very large sample, robust statistical modeling, and recent multi-country DHS data. By contrast, earlier studies often used single-country samples and focused only on the most recent interval, excluding prior, subsequent, and open intervals.

Comparisons with “reverse causation” studies, showing that short interpregnancy intervals elevate child mortality, further illuminate a reinforcing cycle. Meta-analyses and DHS-based reviews consistently report 1.5 to 3-fold higher risks of neonatal, infant, and under-five mortality for intervals <18-24 months, driven by maternal depletion, sibling competition, and infection transmission [9,22,23]. Our findings show how child loss feeds back into this cycle by compressing subsequent intervals, thereby heightening risks for later-born siblings. This bidirectionality has been noted in longitudinal Tanzanian data documenting strong replacement and insurance effects in high-mortality rural settings [18].

This study has several notable strengths that enhance the robustness and generalizability of its findings. First, the use of a large, pooled dataset from recent DHS across 26 sub-Saharan African countries, encompassing over 271,000 women and 1.18 million births, provides greater statistical power and regional representativeness. Second, the application of survey-weighted multilevel Weibull survival models with gamma shared frailty represents a methodological advance, effectively handling right-censoring, complex survey design, intra-woman correlation across repeated intervals, and unobserved maternal heterogeneity. Nevertheless, certain limitations warrant consideration. As with all DHS-based studies, the data are retrospective and rely on maternal recall of birth histories and child survival status, potentially introducing recall bias, particularly for exact dates of births and deaths further in the past. The analysis is restricted to intervals following live births, excluding stillbirths or very early miscarriages that might also trigger replacement behavior.

## Conclusion

The finding that the child replacement effect is widespread across sub-Saharan Africa, as evidenced by this multi-country analysis, reinforces a key policy principle. Gains in child survival must be matched by robust postpartum family planning (PPFP) to support healthy birth intervals and curb accelerated fertility driven by replacement behavior. Programs should integrate bereavement-sensitive PPFP into newborn and child-survival platforms, provide immediate counseling on optimal spacing, ensure access to a full method mix (including LAM) and facilitate timely transition to another method upon menses return [5,24–26]. These actions can help break the cycle in which child mortality compresses intervals, elevates health and mortality risk for subsequent siblings.

## List of abbreviations

AHR: Adjusted Hazard Ratio
CI: Confidence Interval
DHS: Demographic and Health Survey
HR: Hazard Ratio
IQR: Interquartile Range
LAM: Lactational Amenorrhea Method
PPFP: Postpartum Family Planning
SDG: Sustainable Development Goal(s)
SSA: Sub-Saharan Africa
TFR: Total Fertility Rate
WHO: World Health Organization

## Declarations

### Ethical approval and consent to participate

Not applicable

### Consent to publish

Not applicable

### Data availability

The DHS data used for this study are available and can be downloaded upon request at https://dhsprogram.com/data/dataset_admin/login_main.cfm?CFID=386716648&CFTOKEN=41a7550c880afea3-02E3A29E-C13B-B811-8571A7239939AD2F. Accessed on November 7, 2025.

### Authors’ contributions

YM conceived and designed the study, analyzed the data and wrote the manuscript. YM approved the final manuscript.

## Acknowledgments

The DHS data were obtained from the ICF International, Calverton, Maryland, USA.

